## Supplementary material for "Analytic approaches to clinical validation of results from preclinical models of glioblastoma: a systematic review": S1 References

References ordered alphabetically by first author


s1. Alvarado AG, Turaga SM, Sathyan P, et al. Coordination of self-renewal in glioblastoma by integration of adhesion and microRNA signaling. *Neuro-oncology*. 2016;18(5):656-666. doi:10.1093/neuonc/nov196

s2. Bayin N.S., Frenster J.D., Kane J.R., et al. GPR133 (ADGRD1), an adhesion G-protein-coupled receptor, is necessary for glioblastoma growth. *Oncogenesis*. 2016;5(10):e263. doi:10.1038/oncsis.2016.63

s3. Cai H.-Q., Liu A.-S., Zhang M.-J., et al. Identifying Predictive Gene Expression and Signature Related to Temozolomide Sensitivity of Glioblastomas. *Frontiers in Oncology*. 2020;10((Cai, Liu, Liu, Meng, Qian) Department of Neurosurgery, National Cancer Center/National Clinical Research Center for Cancer/Cancer Hospital, Chinese Academy of Medical Sciences and Peking Union Medical College, Beijing, China(Zhang, Wan) Department of Neu):669. doi:10.3389/fonc.2020.00669

s4. Delic S, Lottmann N, Stelzl A, et al. MiR-328 promotes glioma cell invasion via SFRP1-dependent Wnt-signaling activation. *Neuro-oncology*. 2014;16(2):179-190. doi:10.1093/neuonc/not164

s5. Deng S, Li Y, Yi G, et al. Overexpression of COX7A2 is associated with a good prognosis in patients with glioma. *Journal of neuro-oncology*. 2018;136(1):41-50. doi:10.1007/s11060-017-2637-z

s6. Erhart F., Weiss T., Klingenbrunner S., et al. Spheroid glioblastoma culture conditions as antigen source for dendritic cell-based immunotherapy: spheroid proteins are survival-relevant targets but can impair immunogenic interferon gamma production. *Cytotherapy*. 2019;21(6):643-658. doi:10.1016/j.jcyt.2019.03.002

s7. Erhart F, Blauensteiner B, Zirkovits G, et al. Gliomasphere marker combinatorics: multidimensional flow cytometry detects CD44+/CD133+/ITGA6+/CD36+ signature. *Journal of cellular and molecular medicine*. 2019;23(1):281-292. doi:10.1111/jcmm.13927

s8. Genovese G., Ergun A., Shukla S.A., et al. microRNA regulatory network inference identifies miR-34a as a novel regulator of TGF-beta signaling in glioblastoma. *Cancer Discovery*. 2012;2(8):736-749. doi:10.1158/2159-8290.CD-12-0111

s9. Guo R-M, Zhao C-B, Li P, Zhang L, Zang S-H, Yang B. Overexpression of CLEC18B Associates With the Proliferation, Migration, and Prognosis of Glioblastoma. *ASN neuro*. 2018;10(101507115):1759091418781949. doi:10.1177/1759091418781949

s10. Haapa-Paananen S, Chen P, Hellstrom K, et al. Functional profiling of precursor MicroRNAs identifies MicroRNAs essential for glioma proliferation. *PloS one*. 2013;8(4):e60930. doi:10.1371/journal.pone.0060930

s11. Hasan T, Caragher SP, Shireman JM, et al. Interleukin-8/CXCR2 signaling regulates therapy-induced plasticity and enhances tumorigenicity in glioblastoma. *Cell death & disease*. 2019;10(4):292. doi:10.1038/s41419-019-1387-6

s12. Ho K-H, Chen P-H, Hsi E, et al. Identification of IGF-1-enhanced cytokine expressions targeted by miR-181d in glioblastomas via an integrative miRNA/mRNA regulatory network analysis. *Scientific reports*. 2017;7(1):732. doi:10.1038/s41598-017-00826-0 s13. Holmberg Olausson K, Maire CL, Haidar S, et al. Prominin-1 (CD133) defines both stem and non-stem cell populations in CNS development and gliomas. *PloS one*. 2014;9(9):e106694. doi:10.1371/journal.pone.0106694

s14. Hong L, Ya-Wei L, Hai W, et al. MiR-519a functions as a tumor suppressor in glioma by targeting the oncogenic STAT3 pathway. *Journal of neuro-oncology*. 2016;128(1):35-45. doi:10.1007/s11060-016-2095-z

s15. Hu Y, Ylivinkka I, Chen P, et al. Netrin-4 promotes glioblastoma cell proliferation through integrin beta4 signaling. *Neoplasia (New York, NY)*. 2012;14(3):219-227.

s16. Hua T.N.M., Oh J., Kim S., et al. Peroxisome proliferator-activated receptor gamma as a theragnostic target for mesenchymal-type glioblastoma patients. *Experimental and Molecular Medicine*. 2020;52(4):629-642. doi:10.1038/s12276-020-0413-1

s17. Jarboe JS, Anderson JC, Duarte CW, et al. MARCKS regulates growth and radiation sensitivity and is a novel prognostic factor for glioma. *Clinical cancer research : an official journal of the American Association for Cancer Research*. 2012;18(11):3030-3041. doi:10.1158/1078-0432.CCR-11-3091

s18. Kinker GS, Oba-Shinjo SM, Carvalho-Sousa CE, et al. Melatonergic system-based two-gene index is prognostic in human gliomas. *Journal of pineal research*. 2016;60(1):84-94. doi:10.1111/jpi.12293

s19. Klopfenstein Q, Truntzer C, Vincent J, Ghiringhelli F. Cell lines and immune classification of glioblastoma define patient's prognosis. *British journal of cancer*. 2019;120(8):806-814. doi:10.1038/s41416-019-0404-y

s20. Kuang J-Y, Guo Y-F, Chen Y, et al. Connexin 43 C-terminus directly inhibits the hyperphosphorylation of Akt/ERK through protein-protein interactions in glioblastoma. *Cancer science*. 2018;109(8):2611-2622. doi:10.1111/cas.13707

s21. Kudo T., Prentzell M.T., Mohapatra S.R., et al. Constitutive Expression of the Immunosuppressive Tryptophan Dioxygenase TDO2 in Glioblastoma Is Driven by the Transcription Factor C/EBPbeta. *Frontiers in Immunology*. 2020;11((Kudo, Sahm, Platten, Green) DKTK CCU Neuroimmunology and Brain Tumor Immunology, German Cancer Research Center (DKFZ), Heidelberg, Germany(Kudo, Platten, Green) Department of Neurology, Medical Faculty Mannheim, Heidelberg University, Mannheim, Germany(K):657. doi:10.3389/fimmu.2020.00657

s22. Li H, Li J, Chen L, et al. HERC3-Mediated SMAD7 Ubiquitination Degradation Promotes Autophagy-Induced EMT and Chemoresistance in Glioblastoma. *Clinical cancer research : an official journal of the American Association for Cancer Research*. 2019;25(12):3602-3616. doi:10.1158/1078-0432.CCR-18-3791

s23. Li L, Huang Y, Gao Y, et al. EGF/EGFR upregulates and cooperates with Netrin-4 to protect glioblastoma cells from DNA damage-induced senescence. *BMC cancer*. 2018;18(1):1215. doi:10.1186/s12885-018-5056-4

s24. Liu Y, Li X, Zhang Y, et al. An miR-340-5p-macrophage feedback loop modulates the progression and tumor microenvironment of glioblastoma multiforme. *Oncogene*. 2019;38(49):7399-7415. doi:10.1038/s41388-019-0952-x s25. Luedi MM, Singh SK, Mosley JC, et al. A Dexamethasone-regulated Gene Signature Is Prognostic for Poor Survival in Glioblastoma Patients. *Journal of neurosurgical anesthesiology*. 2017;29(1):46-58.

s26. Luedi MM, Singh SK, Mosley JC, et al. Dexamethasone-mediated oncogenicity in vitro and in an animal model of glioblastoma. *Journal of neurosurgery*. 2018;129(6):1446-1455. doi:10.3171/2017.7.JNS17668

s27. Mao D.D., Gujar A.D., Mahlokozera T., et al. A CDC20-APC/SOX2 Signaling Axis Regulates Human Glioblastoma Stem-like Cells. *Cell Reports*. 2015;11(11):1809-1821. doi:10.1016/j.celrep.2015.05.027

s28. Marziali G, Signore M, Buccarelli M, et al. Metabolic/Proteomic Signature Defines Two Glioblastoma Subtypes With Different Clinical Outcome. *Scientific reports*. 2016;6(101563288):21557. doi:10.1038/srep21557

s29. Mega A, Hartmark Nilsen M, Leiss LW, et al. Astrocytes enhance glioblastoma growth. *Glia*. 2020;68(2):316-327. doi:10.1002/glia.23718

s30. Mehrian-Shai R, Yalon M, Simon AJ, et al. High metallothionein predicts poor survival in glioblastoma multiforme. *BMC medical genomics*. 2015;8(101319628):68. doi:10.1186/s12920-015-0137-6

s31. Mikheev AM, Mikheeva SA, Severs LJ, et al. Targeting TWIST1 through loss of function inhibits tumorigenicity of human glioblastoma. *Molecular oncology*. 2018;12(7):1188-1202. doi:10.1002/1878-0261.12320

s32. Nduom EK, Wei J, Yaghi NK, et al. PD-L1 expression and prognostic impact in glioblastoma. *Neuro-oncology*. 2016;18(2):195-205. doi:10.1093/neuonc/nov172

s33. Okura H, Golbourn BJ, Shahzad U, et al. A role for activated Cdc42 in glioblastoma multiforme invasion. *Oncotarget*. 2016;7(35):56958-56975. doi:10.18632/oncotarget.10925

s34. Pandya P., Jethva M., Rubin E., Birnbaum R.Y., Braiman A., Isakov N. PICOT binding to chromatin-associated EED negatively regulates cyclin D2 expression by increasing H3K27me3 at the CCND2 gene promoter. *Cell Death and Disease*. 2019;10(10):685. doi:10.1038/s41419-019-1935-0

s35. Pangeni RP, Zhang Z, Alvarez AA, et al. Genome-wide methylomic and transcriptomic analyses identify subtype-specific epigenetic signatures commonly dysregulated in glioma stem cells and glioblastoma. *Epigenetics.* 2018;13(4):432-448. doi:10.1080/15592294.2018.1469892

s36. Paul Y., Thomas S., Patil V., et al. Genetic landscape of long noncoding RNA (lncRNAs) in glioblastoma: Identification of complex lncRNA regulatory networks and clinically relevant lncRNAs in glioblastoma. *Oncotarget*. 2018;9(51):29548-29564. doi:10.18632/oncotarget.25434

s37. Pollak J, Rai KG, Funk CC, et al. Ion channel expression patterns in glioblastoma stem cells with functional and therapeutic implications for malignancy. *PloS one*. 2017;12(3):e0172884. doi:10.1371/journal.pone.0172884 s38. Polonen P, Jawahar Deen A, Leinonen HM, et al. Nrf2 and SQSTM1/p62 jointly contribute to mesenchymal transition and invasion in glioblastoma. *Oncogene*. 2019;38(50):7473-7490. doi:10.1038/s41388-019-0956-6

s39. Qiu S, Huang D, Yin D, et al. Suppression of tumorigenicity by microRNA-138 through inhibition of EZH2-CDK4/6-pRb-E2F1 signal loop in glioblastoma multiforme. *Biochimica et biophysica acta*. 2013;1832(10):1697-1707. doi:10.1016/j.bbadis.2013.05.015

s40. Rowther FB, Wei W, Dawson TP, et al. Cyclic nucleotide phosphodiesterase-1C (PDE1C) drives cell proliferation, migration and invasion in glioblastoma multiforme cells in vitro. *Molecular carcinogenesis*. 2016;55(3):268-279. doi:10.1002/mc.22276

s41. Safaee M, Clark AJ, Oh MC, et al. Overexpression of CD97 confers an invasive phenotype in glioblastoma cells and is associated with decreased survival of glioblastoma patients. *PloS one*. 2013;8(4):e62765. doi:10.1371/journal.pone.0062765

s42. Sana J, Busek P, Fadrus P, et al. Identification of microRNAs differentially expressed in glioblastoma stem-like cells and their association with patient survival. *Scientific reports*. 2018;8(1):2836. doi:10.1038/s41598-018-20929-6

s43. Sathyan P, Zinn PO, Marisetty AL, et al. Mir-21-Sox2 Axis Delineates Glioblastoma Subtypes with Prognostic Impact. *The Journal of neuroscience : the official journal of the Society for Neuroscience*. 2015;35(45):15097-15112. doi:10.1523/JNEUROSCI.1265-15.2015

s44. Shahar T, Rozovski U, Hess KR, et al. Percentage of mesenchymal stem cells in high-grade glioma tumor samples correlates with patient survival. *Neuro-oncology*. 2017;19(5):660-668. doi:10.1093/neuonc/now239

s45. Sharma A., Bendre A., Mondal A., Muzumdar D., Goel N., Shiras A. Angiogenic gene signature derived from subtype specific cell models segregate proneural and mesenchymal glioblastoma. *Frontiers in Oncology*. 2017;7(JUL):146. doi:10.3389/fonc.2017.00146

s46. Shi Y, Chen C, Yu S-Z, et al. miR-663 Suppresses Oncogenic Function of CXCR4 in Glioblastoma. *Clinical cancer research : an official journal of the American Association for Cancer Research*. 2015;21(17):4004-4013. doi:10.1158/1078-0432.CCR-14-2807

s47. Shugg T., Dave N., Amarh E., Assiri A.A., Pollok K.E., Overholser B.R. Letrozole targets the human ether-a-go-go-related gene potassium current in glioblastoma. *Basic and Clinical Pharmacology and Toxicology*. 2020;((Shugg, Amarh, Assiri, Overholser) Department of Pharmacy Practice, Purdue University College of Pharmacy, West Lafayette, IN, United States(Shugg, Dave, Amarh, Overholser) Division of Clinical Pharmacology, Indiana University School of Medicine, Indianap). doi:10.1111/bcpt.13515

s48. Stegen B, Butz L, Klumpp L, et al. Ca2+-Activated IK K+ Channel Blockade Radiosensitizes Glioblastoma Cells. *Molecular cancer research : MCR*. 2015;13(9):1283-1295. doi:10.1158/1541-7786.MCR-15-0075

s49. Wang C., Zhao N., Zheng Q., Zhang D., Liu Y. BHLHE41 promotes U87 and U251 cell proliferation via ERK/cyclinD1 signaling pathway. *Cancer Management and Research*. 2019;11((Wang, Zhao, Zheng, Zhang, Liu) Department of Pathology, College of Basic Medical Sciences, China Medical University, Shenyang 110122, China(Wang, Zhao,

Zheng, Zhang, Liu) Department of Pathology, The First Affiliated Hospital, China Medical University, S):7657-7672. doi:10.2147/CMAR.S214697

s50. Wen X-M, Luo T, Jiang Y, et al. Zyxin (ZYX) promotes invasion and acts as a biomarker for aggressive phenotypes of human glioblastoma multiforme. *Laboratory investigation; a journal of technical methods and pathology*. 2020;100(6):812-823. doi:10.1038/s41374-019-0368-9

s51. Xavier-Magalhaes A., Goncalves C.S., Fogli A., et al. The long non-coding RNA HOTAIR is transcriptionally activated by HOXA9 and is an independent prognostic marker in patients with malignant glioma. *Oncotarget*. 2018;9(21):15740-15756. doi:10.18632/oncotarget.24597

s52. Xu R, Han M, Xu Y, et al. Coiled-coil domain containing 109B is a HIF1alpha-regulated gene critical for progression of human gliomas. *Journal of translational medicine*. 2017;15(1):165. doi:10.1186/s12967-017-1266-9

s53. Yadav AK, Renfrow JJ, Scholtens DM, et al. Monosomy of chromosome 10 associated with dysregulation of epidermal growth factor signaling in glioblastomas. *JAMA*. 2009;302(3):276-289. doi:10.1001/jama.2009.1022

s54. Yeung YT, Fan S, Lu B, et al. CELF2 suppresses non-small cell lung carcinoma growth by inhibiting the PREX2-PTEN interaction. *Carcinogenesis*. 2020;41(3):377-389. doi:10.1093/carcin/bgz113

s55. Yi G-Z, Xiang W, Feng W-Y, et al. Identification of Key Candidate Proteins and Pathways Associated with Temozolomide Resistance in Glioblastoma Based on Subcellular Proteomics and Bioinformatical Analysis. *BioMed research international*. 2018;2018(101600173):5238760. doi:10.1155/2018/5238760

s56. Zeng A., Yin J., Wang Z., et al. miR-17-5p-CXCL14 axis related transcriptome profile and clinical outcome in diffuse gliomas. *OncoImmunology*. 2018;7(12):e1510277. doi:10.1080/2162402X.2018.1510277

s57. Zhai L, Ladomersky E, Lauing KL, et al. Infiltrating T Cells Increase IDO1 Expression in Glioblastoma and Contribute to Decreased Patient Survival. *Clinical cancer research : an official journal of the American Association for Cancer Research*. 2017;23(21):6650-6660. doi:10.1158/1078-0432.CCR-17-0120

s58. Zhang Y, Cruickshanks N, Yuan F, et al. Targetable T-type Calcium Channels Drive Glioblastoma. *Cancer research*. 2017;77(13):3479-3490. doi:10.1158/0008-5472.CAN-16-2347
