## Supplementary material for "Analytic approaches to clinical validation of results from preclinical models of glioblastoma: a systematic review": S2 Table

**S2 Table**. Refences to specific analyses extracted for comparison of results on molecular markers

| Molecular marker | Author | Analysis type | Association | Location of analysis in manuscript |
|---|---|---|---|---|
| CXCL14 | Zeng 2018 | U | ▼ | Figure 3K reporting p=0.02809 |
| | | M | □ | Supplementary Table 2 reporting HR 1.04 p=0.352 |
| EGFR | Kuang 2018 | U | ▼ | Figure 7, top survival curve in left panel reporting p=0.0009 |
| | Li 2018 | U | □ | Figure 4A lower panel reporting p=0.216 |
| HOTAIR | Xavier-Magalhaes 2018 | U | ▼ | Figure 4B p=0.032 |
| | | M | ▼ | Supplementary Table 2 p=0.036 |
| IDO1 | Zhai 2017 | U | ▼ | Figure 2C right panel, p<0.05 |
| | | M | ▼ | Table 1, HR 1.82 (95% CI 1.17-2.81) p=0.0076 |
| IL-8 | Hasan 2019 | U | ▼ | Figure 2B top left panel p=0.0112 |
| | | M | ▼ | Legend of Figure 2B, HR 1.07, 95% CI 1.01-1.14), p=0.0467 |
| MARCKS | Jarboe 2012 | U | ▲ | Figure 6B, p=0.0002 |
| | | M | ▲ | Supplementary Table 2, HR 0.21 (95% CI 0.09-0.48), p=0.0002 |
| miR-17-5p | Zeng 2018 | U | ▲ | Figure 6D reporting p=0.0006 |
| | | M | □ | “After incorporating the age and molecular subtype factors, the analysis indicated that miR-17-5p value was not an independent prognostic marker for the overall survival of GBM patients” |
| miR-181d | Genovese 2012 | U | □ | Supplementary Table 6 on page 606 of supplementary PDF file, p=0.46 |
| | Ho 2017 | U | □ | Figure 4D, p=0.067 |
| miR-34a | Genovese 2012 | U | ▼ | Figure 3D reporting p=0.0154 |
| | | M | ▼ | Supplementary Table 7 on page 607 of supplementary PDF file reporting p=0.0016 |
| NTN4 | Hu 2012 | U | ▲ | Figure 3B, p<0.05 |
| | Li 2018 | U | □ | Figure 4B, lower panel, p=0.753 |
| PD-L1 | Nduom 2016 | U | ▼ | Figure 4A, p=0.231 |
| | | M | ▼ | Text: “HR 1.54, 95% CI 1.05-2.28, p=0.0231” |
| POSTN | Mega 2020 | U | ▼ | Table 2, HR 1.35, 95% CI 1.11-1.64, p=0.002 |
| | Liu 2019 | U | ▼ | Figure 7h, p=0.0003 |
| | Mega 2020 | M | ▼ | Table 2, HR 1.37, 95% CI 1.06-1.77, p=0.017 |
| SFRP1 | Delic 2014 | U | ▲ | Figure 7B, p<0.05 |
| | | M | ▲ | Table 1, HR 0.782 (95%CI 0.642-0.953), p=0.015 |
| Sox2 | Sathyan 2015 | U | ▲ | Figure 6C, 0=0.0307 |
| | | M | □ | Text “…after correcting for age and KPS, Sox2 lost significance (p=0.0964)” |
| SRGN | Mega 2020 | U | ▼ | Table 2, HR 1.63, 95% CI 1.19-2.23, p=0.002 |
| | | M | □ | Table 2, HR 1.29, 95% CI 0.83-2.00, p=0.263 |

Molecular markers ordered alphabetically accompanied with the location of analysis in the original manuscript. U = univariable survival analysis; M = multivariable survival analysis; ▲ = positive association i.e. higher levels of the molecular marker associated with better survival and p<0.05; ▼= negative association i.e. lower levels of molecular marker associated with worse survival and p<0.05; □ = statistical significance not demonstrated (p≥0.05)
