## Supplementary material for "Analytic approaches to clinical validation of results from preclinical models of glioblastoma: a systematic review": S1 Table

**S1 Table.** Characteristics of 58 included studies

| Author, year of publication | Cohort selection described | Data source [number of patients] | | Data type | Molecular marker(s) or set of variables of interest | | | | Survival analysis | |
|---|---|---|---|---|---|---|---|---|---|---|
| | | TCGA [number] | CGGA [number] | | One | More than one | Set of markers | Set of markers and clinical variables | Univariable | Multivariable |
| Alvarado, 2016 | No | Yes [NS] | No [--] | B | No | Yes | Yes | No | Yes | No |
| Bayin, 2016 | No | Yes [160] | No [--] | C | Yes | No | No | No | Yes | No |
| Cai, 2020 | Yes | Yes [103] | Yes [83] | C | No | No | Yes | Yes | Yes | Yes |
| Delic, 2014 | No | Yes [519] | No [--] | B | No | Yes | No | No | Yes | Yes |
| Deng, 2017 | No | Yes [540] | No [--] | A | Yes | No | No | No | Yes | Yes |
| Erhart, 2018 | No | Yes [1022] | No [--] | B | No | No | Yes | No | Yes | No |
| Erhart, 2019 | No | Yes [660] | No [--] | A | No | No | Yes | No | Yes | No |
| Genovese, 2012 | No | Yes [290] | No [--] | B | No | Yes | No | No | Yes | No |
| Guo, 2018 | No | Yes [169] | No [--] | A | Yes | No | No | No | Yes | Yes |
| Haapa-Paananen, 2013 | No | Yes [308] | No [--] | B | No | Yes | No | No | Yes | No |
| Hasan, 2019 | No | Yes [265] | No [--] | A | Yes | No | No | Yes | Yes | Yes |
| Ho, 2017 | No | Yes [694] | No [--] | F | No | Yes | Yes | No | Yes | Yes |
| Holmberg Olausson, 2014 | No | Yes [NS] | No [--] | A | Yes | No | No | No | Yes | No |
| Hong, 2016 | Yes | Yes [170] | No [--] | E | Yes | No | No | Yes | Yes | No |
| Hu, 2012 | No | Yes [425] | No [--] | A | No | Yes | No | No | Yes | No |
| Hua, 2020 | No | Yes [206] | No [--] | A | Yes | No | No | No | Yes | No |
| Jarboe, 2012 | Yes | Yes [192] | No [--] | H | Yes | No | No | Yes | Yes | Yes |
| Kinker, 2016 | No | Yes [152] | No [--] | C | Yes | No | Yes | No | Yes | Yes |
| Klopfenstein, 2019 | Yes | Yes [481] | No [--] | A | No | Yes | Yes | No | Yes | Yes |
| Kuang, 2018 | No | Yes [540] | No [--] | A | Yes | No | Yes | No | Yes | No |
| Kudo, 2020 | No | Yes [525] | No [--] | A | No | Yes | No | No | Yes | Yes |
| Li, 2018 | Yes | Yes [92] | No [--] | C | No | Yes | Yes | No | Yes | No |
| Li, 2019 | No | Yes [NS] | No [--] | D | Yes | No | No | No | Yes | No |
| Liu, 2019 | No | Yes [565] | No [--] | B | No | Yes | No | No | Yes | Yes |
| Luedi, 2017 | No | Yes [515] | No [--] | A | No | No | Yes | No | Yes | Yes |

| | | | | | | | | | | |
|---|---|---|---|---|---|---|---|---|---|---|
| Luedi, 2018 | No | Yes [520] | No [--] | A | No | No | Yes | No | Yes | No |
| Mao, 2015 | No | Yes [473] | No [--] | A | Yes | No | No | No | Yes | Yes |
| Marziali, 2016 | No | Yes [251] | No [--] | C | No | No | Yes | No | Yes | Yes |
| Mega, 2020 | Yes | Yes [569] | No [--] | A | No | Yes | Yes | No | Yes | Yes |
| Mehrian-Shai, 2015 | Yes | Yes [210] | No [--] | A | No | Yes | No | No | Yes | No |
| Mikheev, 2018 | No | Yes [525] | No [--] | A | Yes | No | No | No | Yes | No |
| Nduom, 2016 | No | Yes [149] | No [--] | C | No | Yes | Yes | No | Yes | Yes |
| Okura, 2016 | Yes | Yes [528] | No [--] | A | Yes | No | No | No | Yes | No |
| Polonen, 2019 | No | Yes [520] | No [--] | D | Yes | No | No | No | Yes | Yes |
| Pandya, 2019 | No | Yes [142] | No [--] | A | No | Yes | No | No | Yes | No |
| Pangeni, 2018 | Yes | Yes [348] | No [--] | A | Yes | No | No | No | Yes | No |
| Paul, 2018 | Yes | Yes [172] | No [--] | G | No | Yes | Yes | No | Yes | Yes |
| Pollak, 2017 | Yes | Yes [525] | No [--] | A | No | Yes | No | No | Yes | No |
| Qiu, 2013 | Yes | Yes [480] | No [--] | B | Yes | No | No | No | Yes | No |
| Rowther, 2016 | No | Yes [596] | No [--] | A | No | No | Yes | No | Yes | No |
| Safaee, 2013 | No | Yes [212] | No [--] | A | Yes | No | No | No | Yes | No |
| Sana, 2018 | Yes | Yes [485] | No [--] | E | No | No | Yes | No | Yes | Yes |
| Sathyan, 2015 | Yes | Yes [353] | No [--] | B | No | Yes | Yes | No | Yes | Yes |
| Shahar, 2017 | No | Yes [414] | No [--] | A | No | Yes | Yes | No | Yes | Yes |
| ShaRNA, 2017 | No | Yes [39] | No [--] | D | No | Yes | No | No | Yes | No |
| Shi, 2015 | No | Yes [476] | No [--] | B | No | Yes | Yes | No | Yes | Yes |
| Shugg, 2020 | Yes | Yes [152] | No [--] | A | No | Yes | No | No | Yes | No |
| Stegen, 2015 | No | Yes [NS] | No [--] | C | Yes | No | No | No | Yes | No |
| Wang, 2019 | No | Yes [153] | No [--] | A | Yes | No | No | No | Yes | No |
| Wen, 2020 | No | Yes [264] | No [--] | A | Yes | No | No | No | Yes | No |
| Xavier-Magalhaes, 2018 | Yes | Yes [554] | No [--] | I | Yes | No | No | Yes | Yes | Yes |
| Xu, 2017 | No | Yes [NS] | Yes [NS] | A | Yes | No | No | No | Yes | Yes |
| Yadav, 2009 | No | Yes [407] | No [--] | B | Yes | No | No | No | Yes | Yes |
| Yeung, 2020 | No | Yes [573] | No [--] | J | Yes | No | No | No | Yes | No |

| | | | | | | | | | | |
|---|---|---|---|---|---|---|---|---|---|---|
| Yi, 2018 | No | Yes [540] | No [--] | J | No | Yes | No | No | Yes | No |
| Zeng, 2018 | No | Yes [NS] | No [--] | F | No | Yes | No | No | Yes | Yes |
| Zhai, 2017 | No | Yes [172] | No [--] | D | Yes | No | No | No | Yes | Yes |
| Zhang, 2017 | No | Yes [607] | No [--] | J | Yes | No | No | No | Yes | No |

Data type:

A = RNA microarray only

B = RNA microarray and miRNA microarray

C = RNA sequencing only

D = RNA microarray and RNA sequencing

E = miRNA microarray only

F = RNA sequencing, RNA microarray and miRNA microarray

G = RNA sequencing and miRNA microarray

H = RNA microarray and DNA methylation

I = RNA sequencing, RNA microarray and DNA methylation

J = Unspecified

If a study used a data source but not specified the number of patients, the column for data source would be “Yes [NS]” indicating number of patients not specified.
