## Supplementary figures and images for "Analytic approaches to clinical validation of results from preclinical models of glioblastoma: a systematic review"

### S1 Fig

**S1 Figure. Common analytic strategy used by included studies.**

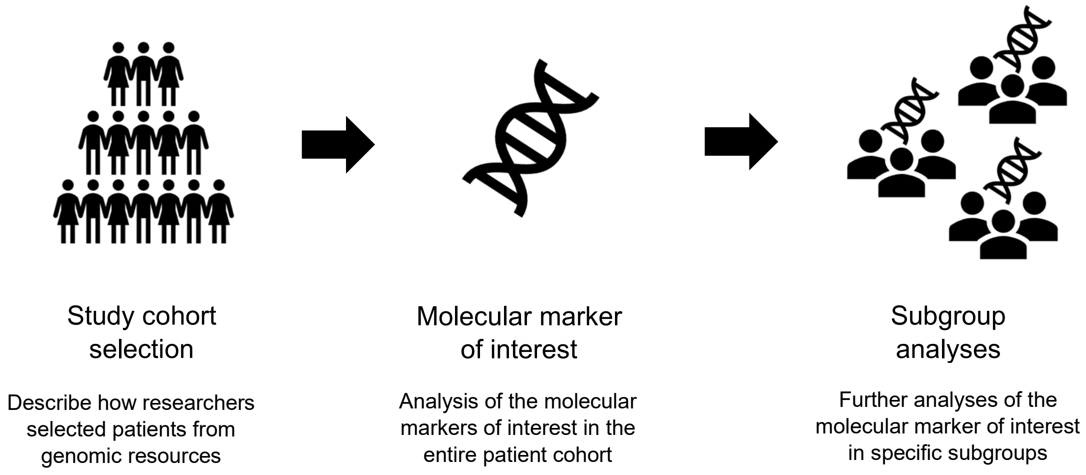
