## Supplementary material for "Analytic approaches to clinical validation of results from preclinical models of glioblastoma: a systematic review": S2 Supporting Information

Search strategy

*Medline:*

1. (glioblastoma or gbm or (grade adj2 IV adj2 glioma) or (grade adj2 4 adj2 glioma) or (glioblastoma adj2 multiforme) or glioma*).mp or exp glioblastoma/ or exp glioma/
2. (surviv* or outcome or prognos* or mortality).mp or exp mortality/ or exp survival/ or exp prognosis/
3. (predict* or associa* or scor*).mp
4. (TCGA or (the cancer genome atlas) or CGGA or (Chinese glioma genome atlas)).mp
5. review/ or editorial/
6. and/ 1-4 not 5
7. limit 6 to yr= “2008 -Current”

*Embase*:

1. (glioblastoma or gbm or (grade adj2 IV adj2 glioma) or (grade adj2 4 adj2 glioma) or (glioblastoma adj2 multiforme) or glioma*).mp or exp glioblastoma/ or exp glioma/
2. (surviv* or outcome or prognos* or mortality).mp or exp mortality/ or exp survival/ or exp prognosis/ or exp cancer prognosis/
3. (predict* or associa* or scor*).mp
4. (TCGA or (the cancer genome atlas) or CGGA or (Chinese glioma genome atlas)).mp
5. review/ or editorial/
6. and/ 1-4 not 5
7. limit 6 to yr=”2008 -Current”
