## Supplementary material for "Analytic approaches to clinical validation of results from preclinical models of glioblastoma: a systematic review": S1 Supporting Information

Eligibility criteria

Inclusion:

- Patients (≥ 18 years old) diagnosed with non-recurrent histopathologically confirmed glioblastoma according to WHO classification
- Studies which utilised TCGA/CGGA resources or included TCGA/CGGA patients with other patients
- Studies reporting overall survival
- Studies reporting on association between genetic or molecular markers with survival
    - Gene expression, variants, methylation

Exclusion:

- Case reports, reviews, editorials
- Studies which only report their own patient cohort and does not include TCGA/CGGA resources
- Studies which report results only for patients (< 18 years old)
- Studies which include patients < 18 years old and have not separated their data from those >18
- Studies that reported progression free survival only
- Studies including patients with recurrent glioblastoma only
- Patients without a histopathological confirmed glioblastoma diagnosis according to WHO classification
